# Behavioral Markers for Deficits in Speed of Processing in Cerebrovascular Disease

**DOI:** 10.1101/2021.09.23.21263859

**Authors:** Ying Chen, Kelly M Sunderland, Yuelee Khoo, Paula M. McLaughlin, Donna Kwan, Julia Fraser, Joel Ramirez, Malcolm A. Binns, Stephen R. Arnott, Derek Beaton, Donald C. Brien, Leanne K. Casaubon, Brian C. Coe, Benjamin Cornish, Dariush Dowlatshahi, Ayman Hassan, Brian Levine, Wendy Lou, Jennifer Mandzia, William McIlroy, Manuel Montero-Odasso, Karen Van Ooteghem, Joseph B. Orange, Alicia J. Peltsch, Frederico Pieruccini-Faria, Pradeep Reddy Raamana, Angela C. Roberts, Demetrios Sahlas, Gustavo Saposnik, Stephen C. Strother, Richard H. Swartz, Angela K. Troyer, the ONDRI Investigators, Douglas P. Munoz

## Abstract

**Objective:** To assess overlap and uniqueness of established behavioral markers of speed of processing for different aspects of visual information within a cerebrovascular disease cohort, and to examine the link between these speed of processing markers and functional behavior, specifically walking.

**Methods:** A cohort of 161 participants with cerebrovascular disease recruited to the Ontario Neurodegenerative Disease Research Initiative (ONDRI) were examined with three types of assessments: neuropsychology, saccadic eye movement and gait. Principal component analysis (PCA) and canonical correlation analysis (CCA) were performed on select variables from these assessments to reveal commonalities and discrepancies among the measures.

**Results:** PCA analysis revealed different variable patterns between neuropsychology and saccade assessments, with the first component characterized primarily by neuropsychology, and the second and third components more influenced by the saccade assessments. CCA analysis did not reveal association between different types of assessments with the exception of a modest, but significant, positive association between speed of processing measures from the neuropsychological assessments and gait speed.

**Discussion:** Neuropsychological tests and the pro-saccade task can be used for assessment of speed of processing for two major features of visual information, visual perception vs. spatial location. Despite a general lack of association between different types of assessments, combining gait speed as an important contributor to the models reinforces the idea of the link between speed of processing and complex function such as walking, and provides support for the importance of attending to the potential consequences of changes in speed of processing after neurologic injury.

## 1. INTRODUCTION

Impairment in a broad range of cognitive domains is often observed in individuals following an ischemic stroke^1-6^. Fundamental deficits in speed of processing can underlie impairments in multiple cognitive domains; as such, improving processing speed has been suggested as a target for early intervention in individuals after stroke^7^. Most individuals recovering from stroke report that slowed information processing limited activities of daily living^8^. Interestingly, speed of processing contributes to functional outcomes independent of other factors (e.g., age, gender, education, depression)^9^.

Speed of processing is assessed clinically using behavioral tasks that rely on a response-to-sensory input. Such assessments often involve vision-dependent behaviors including commonly used neuropsychological tests such as the Symbol Digit Modalities Test (SDMT)^10-12^ or visually guided movement tasks such as saccadic eye movement^13, 14^. In the former, task performance relies on visual perception of numbers and digits, while saccadic eye movement depends on visual spatial ability. Functional tasks such as walking have demonstrated links with direct markers of speed of processing including saccadic eye movement^15^, and indirect markers including reaction time to verbally responding to an auditory stimulus^16^. While gait speed is influenced by a complex set of factors in stroke, including motor impairments and stroke severity, it remains possible that impairment in gait speed during preferred and/or fast paced walking may be partially explained by deficits in speed of processing.

Most often, a single type of behavioral task is used to assess speed of processing in individuals following stroke^17-22^, despite the possibility that deficits in speed of processing may be differentially revealed by task type, by contrast in performance across tasks, and/or linked to functional behavior. The ability to detect their relationship in the phase of mild to moderate stroke recovery, as well as their potential link to functional tasks, serves as important foundational work for understanding interactions across the recovery continuum.

We investigated two different types of behavioral assessments that directly measure speed of processing including neuropsychological tests and saccadic eye movements in a cerebrovascular disease cohort recruited as part of the Ontario Neurodegenerative Disease Research Initiative (ONDRI)^23, 24^. This was extended to examine the relationship of the clinical measures of speed of processing to gait performance, specifically speed of walking. Important to this study was exploration of speed of processing from multiple types of behavioral tasks, including standard neuropsychological tests and saccadic eye movements, and a functional behavior, gait. We hypothesized that (1) deficits in speed of processing for different aspects of visual information, and (2) distinct cross-assessment relationships would be revealed by combining measures from the three types of behavioral tasks. The potential that a cross-modal index of speed of processing could distinguish individuals within this cohort, unique from unimodal assessment, would reinforce the idea that speed of processing may be a fundamental function that may impact a broad range of control systems and behaviors worthy of unique attention when assessing change after neurological injury.

## 2. METHOD

### 2.1 Participants

We used baseline data from participants with cerebrovascular disease (CVD) enrolled in the ONDRI study. The ONDRI study is a multi-site, longitudinal project with four neurodegenerative disease cohorts and a cerebrovascular disease cohort, collecting data from multiple assessment platforms^24^. Detailed inclusion and exclusion criteria for the CVD cohort are described previously^23,24^. Briefly, CVD participants had experienced an ischemic stroke event documented on MRI or CT at least three months prior to enrollment. Silent strokes (as seen on CT or MRI but without clinical history of focal neurological deficits) were included. Participants with no other vascular cause of symptoms (e.g. migraine, isolated vertigo), those with large cortical strokes (>1/3 middle cerebral artery), and those with severe cognitive impairment, aphasia, or functional disability limiting their ability to complete the study protocol were excluded. Ethics approval was obtained from all participating institutions. The participants provided their written informed consent.

### 2.2 Behavioral tasks and variables

Neuropsychology: The detailed neuropsychology protocol was described previously^25^. For this study, we included two conditions of the Delis Kaplan Executive Function System (DKEFS) Color Word Interference Test (color naming trial and word reading trial), SDMT, and Trail Making Test (TMT) part A. These four tests have been commonly used in measuring speed of processing both clinically and in research^26-30^. For the two DKEFS conditions, participants were asked to name the color of squares, or read the words “red”, “green”, and “blue”^31^. In the SDMT test, participants were asked to write digits matching the nine abstract symbols in 90 seconds according to a given key of digit-symbol pairs^32^. An oral version was administered when participants were not able to write. In the TMT part A test, participants were required to draw a line connecting 25 consecutive numbers arranged pseudo-randomly^33^. We included the following variables to assess speed of processing: completion time for each of the two DKEFS trials (DKEFS-CN, DKEFS-WR, respectively), total number of correct responses in SDMT (SDMT_Total), and completion time for TMT part A (TMT-A).

Saccadic eye movement: Participants performed pro- and anti-saccade tasks that were pseudo-randomly interleaved, with 120 trials in each of two blocks. Each trial began with the appearance of a central fixation point either in green or red, indicating a pro- or anti-saccade task, respectively, followed by a gap period of 200ms with the fixation being removed and screen becoming blank. A peripheral stimulus then appeared 10° left or right horizontally to the original central fixation. On each pro-saccade trial, participants were required to make a saccade to the stimulus location as quickly as possible when it appeared. An infrared camera-based eye tracker (Eyelink 1000 Plus; SR Research Ltd, Ottawa, ON, Canada) was applied to track monocular eye position at a sampling rate of 500 Hz. To assess processing speed, median pro-saccade regular reaction time (PS_Reg_RT) on correct trials, and percentage of express latency pro-saccades (PS_per_exp) were considered, where saccadic reaction time was measured as the time from peripheral stimulus appearance to the onset of the first saccade towards it. Saccades with reaction time of 140-800ms were defined as regular latency saccade, and those with reaction time of 90-139ms were termed as express latency saccade^34^.

Gait: The gait and balance protocol was described previously^35^. Average gait speed over three preferred walking trials (PW_spd), and gait speed from one fast walking trial (FW_spd) were examined. For the preferred gait task, participants walked at their usual pace. For the fast walking task, participants were instructed to walk as fast and safely as they could. For the purpose of this study, these two gait speed variables were included as indirect measures of speed of processing; drawing on evidence of the association between gait speed and speed of processing in older adults^36^ and stroke^37^, which suggests some shared central nervous system control^15^.

### 2.3. Statistical analyses

R software (version 3.6.1) was used for statistical analyses. In addition to the eight selected speed of processing variables, we examined demographic, clinical, and global cognition variables for the cohort. Reaction time variables (DKEFS_CN, DKEFS_WR, TMT-A, PS_Reg_RT) were reverse-coded so that a higher score represented better performance across all variables. Principal component analysis (PCA)^38^ and canonical correlation analysis (CCA)^39^ were used to identify the unique and overlapping aspects of speed of processing cross-assessment measures. To adjust for the effects of age, sex and education level, each variable was first fit with a linear regression model including the three potential confounders as main effects. Residuals of the models were extracted, standardized, and subsequently used as the observed values in PCA and CCA.

Variable patterns within each selected PCA component were represented by component loadings and examined. Contribution of each variable was the squared loading divided by the eigenvalue of the component. Total contribution of all variables within each type of assessment was the sum of contributions from individual variables. The Montreal Cognitive Assessment (MoCA) was used to evaluate global cognition; its relationship with speed of processing was assessed by correlation analysis between PCA component scores and total MoCA scores. Wilk’s λ test was used to estimate significance (p < 0.05) of the canonical functions. For individual functions that reached significance, canonical variate correlation coefficients (Rc) were examined, and correlations between each variable and their canonical variate were reported using canonical coefficients (CCoef) and structure correlations (r_s_).

Mechanisms for missing data were assessed with observed missing codes where data were missing at random. Imputation was performed with regularized iterative PCA^40^ using the missMDA package in R^41^ so that missing data were imputed based on the relative performance of the participant across the behavioral tasks. When a participant was missing all data in two or more types of tasks, they were excluded from the study, as we did not feel there was sufficient data available for imputation.

## 3. RESULTS

### 3.1. Participant characteristics

ONDRI’s CVD cohort includes 161 participants. One participant did not provide sufficient usable data for saccades and could not perform the gait assessments, thus was consequently excluded from analysis. There was a small proportion of missing data (2%; 30 missing values over the total of 1280 data points) within the remaining 160 participants. In particular, one value was missing across the neuropsychology assessment, 16 values were missing for the pro-saccade variables, and 13 values were missing among the gait variables. All missing data were assumed missing at random based on observed missing codes. Characteristics of the 160 participants are summarized in Table 1. Of the 160 participants, 74 (46%) individuals had cognitive impairment according to the MoCA threshold of 26^42^.

**Table 1.**
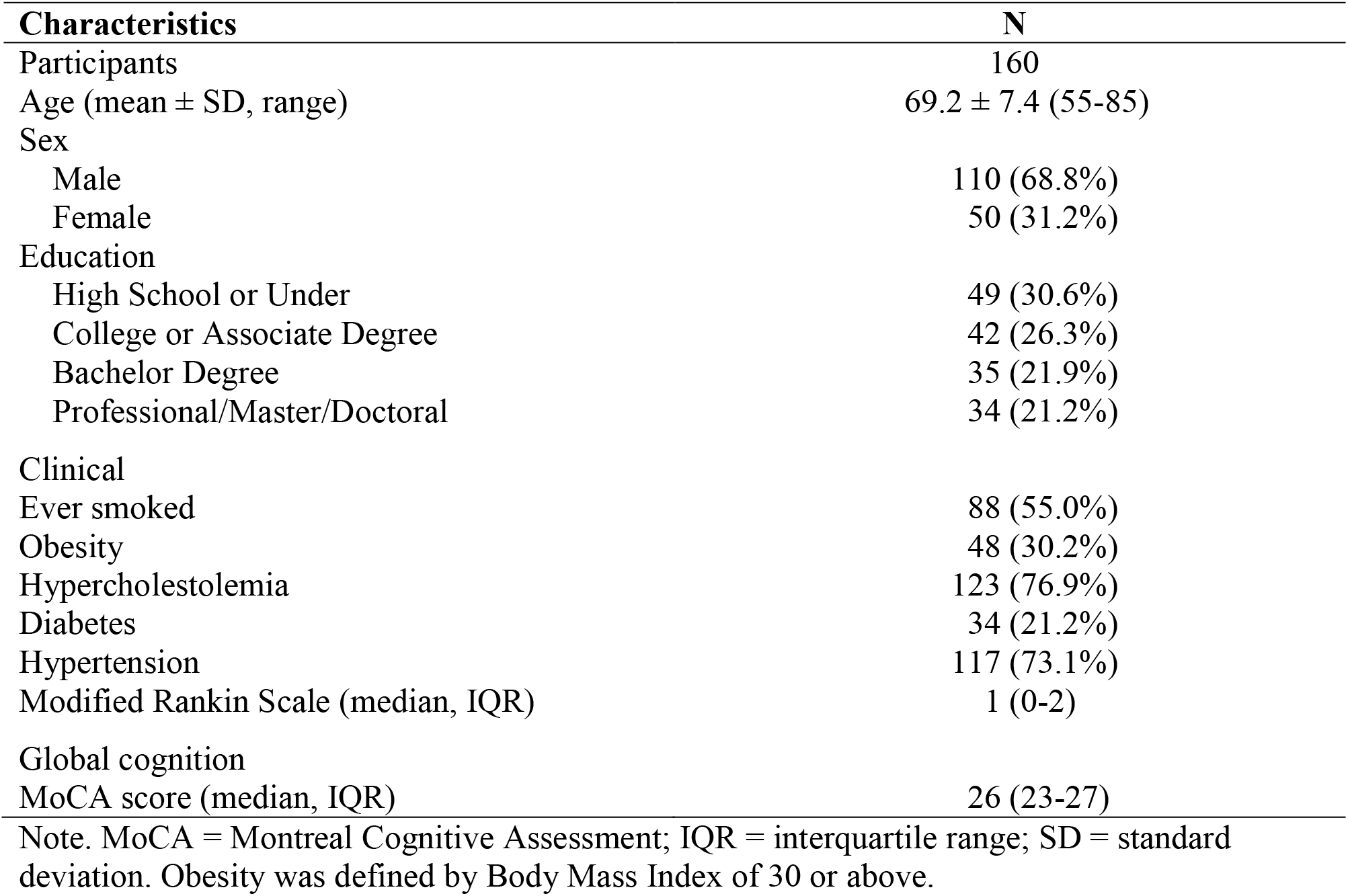
Characteristics of participants

### 3.2. Principal Component Analysis (PCA)

PCA was used to examine patterns within this population among the speed of processing variables. The first three components explained 74.9% of variance in the data (35.4%, 22.2%, and 17.3%, respectively) and were retained for interpretation. Component loadings from individual variables are plotted in Fig. 1, and show that variables within each assessment type are highly correlated. Corresponding component loadings and total contributions within each type of assessment are reported in Table 2. The first component is largely characterized by the four neuropsychology variables, while the second and third components are characterized by the pro-saccade variables, along with the gait variables having the similarities or differences from the pro-saccade variables for second and third component, respectively.

**Table 2.** Component loadings of variables and overall contributions from each of the three types of tasks to components 1 to 3.

| Tasks | Variables | 1<sup>st</sup> Component |  | 2<sup>nd</sup> Component |  | 3<sup>rd</sup> Component |  |
| --- | --- | --- | --- | --- | --- | --- | --- |
|  |  | Loadings | Contributions | Loadings | Contributions | Loadings | Contributions |
| NPSY | DKEFS_CN | 0.822 |  | -0.183 |  | 0.134 |  |
|  | DKEFS_WR | 0.823 | 0.839 | -0.232 | 0.086 | 0.164 | 0.079 |
|  | SDMT_Total | 0.726 |  | -0.075 |  | 0.199 |  |
|  | TMT-A | 0.700 |  | -0.244 |  | 0.156 |  |
| EYE | PS_Reg_RT | 0.024 | 0.006 | 0.574 | 0.400 | 0.668 | 0.592 |
|  | PS_per_exp | -0.122 |  | 0.619 |  | 0.609 |  |
| GAIT | PW_spd | 0.518 | 0.156 | 0.658 | 0.514 | -0.450 | 0.330 |
|  | FW_spd | 0.414 |  | 0.694 |  | -0.503 |  |

**Figure 1.**
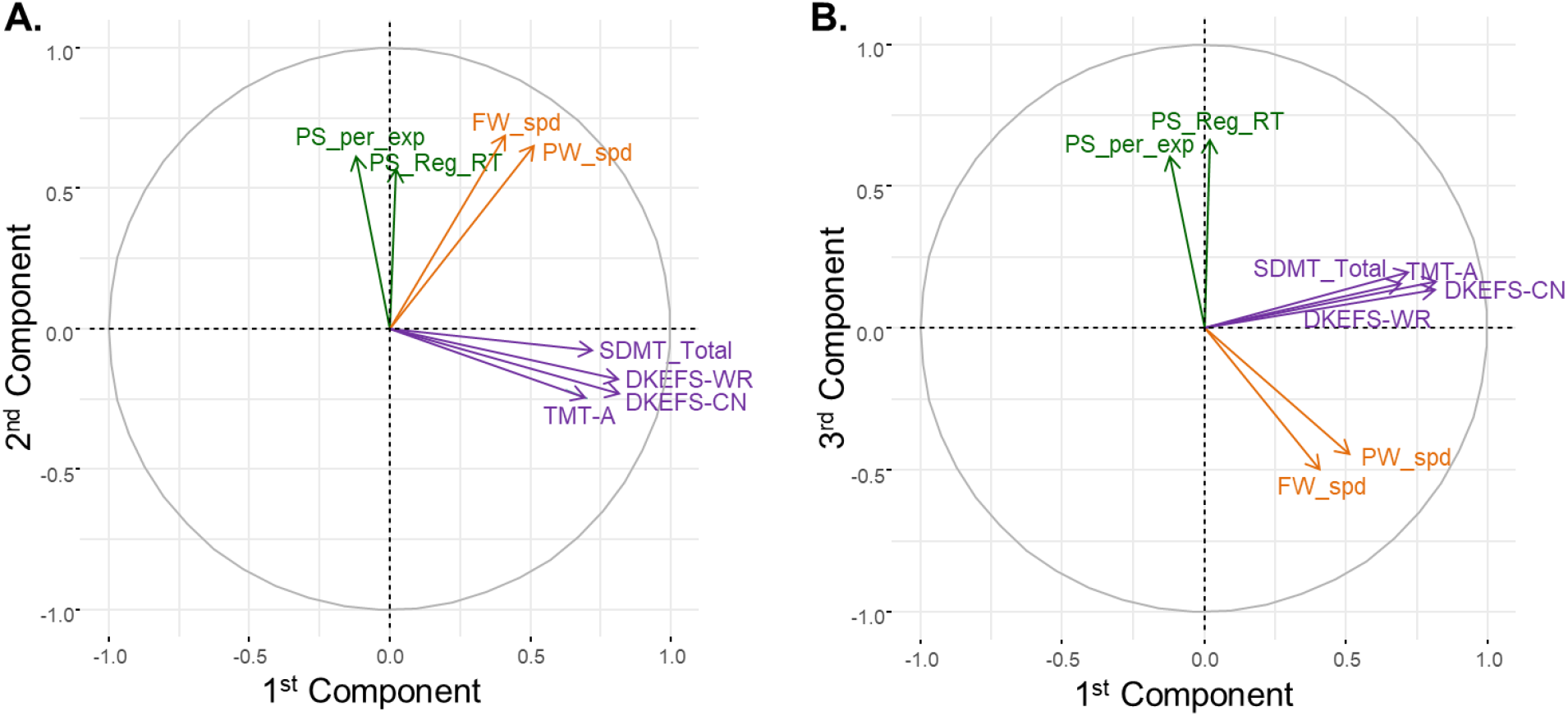
PCA Component loadings of variables on the first three components. A) First and second components. B) First and third components.

The correlation between total MoCA score the first component was r = 0.45 (p<0.001). The correlation between the MoCA and other components was not significant.

### 3.3. Canonical Correlation Analysis (CCA)

The CCA was used to identify associations between each pair of behavioral assessment types. Thus, three CCAs were conducted (Table 3). Wilk’s λ test showed that only the full neuropsychology-gait model was statistically significant (λ = 0.882, F (8, 308) = 2.49, p = 0.01), with 11.8% of the shared variance between gait and neuropsychology measures explained. Within the full model, only the first canonical function was significant (R_c_ = 0.327). Further, as shown in Table 4, DKEFS-WR from neuropsychology and PW_spd from gait had the largest correlation with their corresponding variates.

**Table 3.** Significance test of canonical functions.

| Canonical functions | Wilk's $\lambda$ | F | Hypo. df | Error df | p |
| --- | --- | --- | --- | --- | --- |
| Neuropsychology + Gait |  |  |  |  |  |
| Both functions | 0.8820 | 2.49 | 8 | 308 | 0.01 |
| 2<sup>nd</sup> function | 0.9872 | 0.67 | 3 | 155 | 0.57 |
| Neuropsychology + Eye movement |  |  |  |  |  |
| Both functions | 0.9594 | 0.81 | 8 | 308 | 0.60 |
| 2<sup>nd</sup> function | 0.9889 | 0.58 | 3 | 155 | 0.63 |
| Gait + Eye movement |  |  |  |  |  |
| Both functions | 0.9903 | 0.38 | 4 | 312 | 0.82 |
| 2<sup>nd</sup> function | 0.9995 | 0.07 | 1 | 157 | 0.79 |

Note. Hypo = hypothesis.

**Table 4.**
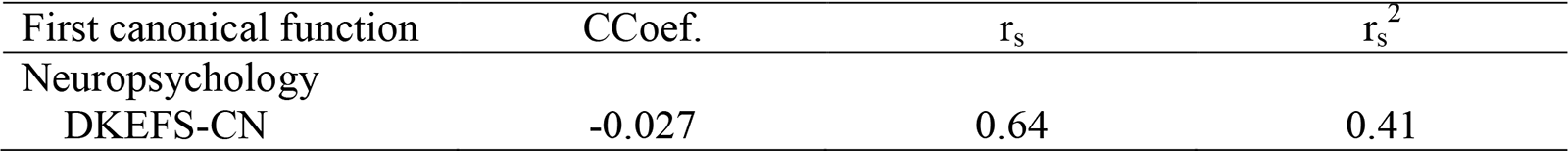
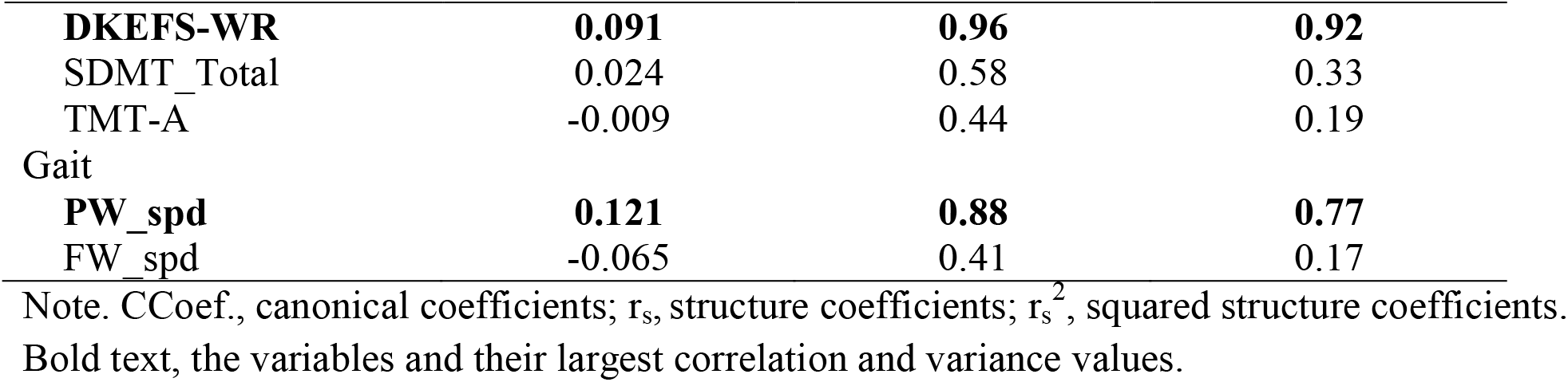
First function of canonical variates for neuropsychology and gait variable sets.

## 4. DISCUSSION

This study used a multi-modal approach to examine measures from two different types of behavioral assessments that probe deficits in speed of processing directly, including neuropsychology and saccadic eye movements, and explored their relationship to a functional behavior, namely gait, in 160 participants with cerebrovascular disease. We found that measures of speed of processing from the four neuropsychological tests versus those from the pro-saccade task share few commonalities, while a modest positive association between neuropsychological assessment and gait speed was present.

The principal component analysis revealed task-related differences; specifically, neuropsychological variables loaded heavily onto the first component, whereas pro-saccade variables loaded more heavily onto the second and third components. One explanation for the different loadings is that respective tasks required distinct aspects of visual information processing: the neuropsychological tests required visual perception of numbers, symbols or colors, whereas the pro-saccade task required visual processing of stimulus spatial location. Given this difference in task demands, measures from neuropsychology and pro-saccade may be considered distinct behavioral markers for visual perception and spatial location^43, 44^subtypes of speed of processing, respectively. This can be further supported by the two visual streams model where both streams arise from primary visual cortex, but the dorsal stream extending into the posterior parietal cortex is involved in processing of spatial location of a target for goal-driven action such as saccade, whereas the ventral stream projecting to the infero-temporal cortex is associated with visual perception of intrinsic characteristics such as size and form^45, 46^.

It has been argued that deficits in speed of processing should be included in the post-stroke cognitive profile as a domain^47^. Our findings provide insight into what behavioral tasks could be used, which may in turn focus development of rehabilitation strategies. Considering a target on speed of processing for visual perception, neuropsychology tests would be selected for assessment. On the other hand, a pro-saccade task would be considered for assessment and training purpose for deficits in speed of processing for spatial localization.

The gait speed variables had heavy loadings on both first and second components, suggesting that gait speed may be impacted by speed of processing. The significant relationship between neuropsychology and gait revealed by CCA analysis may be interpreted by sensorimotor processing demands that impact gait speed. This, together with the observed relationship between gait speed and global cognition in another study with participants following ischemic stroke^48^, suggests that gait speed as a whole may serve as a global and sensitive indicator of impairment rather than specific elements of cognition. In addition, the modest degree of association demonstrates the complexity of this relationship (i.e., the interplay between motor and cognitive function) which is further highlighted by the finding that preferred gait speed had the largest contribution in the canonical function. It may be that fast gait speed, compared to preferred walking speed, is more heavily influenced by factors unrelated to speed of processing such as muscle power or even balance confidence.

The loadings of pro-saccade and gait variables on opposite sides in the third principal component may be related to the two orienting attention systems^49^, a more dorsal pathway interacting with the processing of a stimulus location for goal-directed movement versus a ventral pathway being involved in orienting to objects in a visual scene. The significant correlation between total MoCA score and score of first principal component mainly consisting of the neuropsychological outcomes was not a surprise because we know that the MoCA and the four neuropsychological tests have many characteristics in common. There was no relationship between total MoCA score and score of either second or third principal component, suggesting that the MoCA as a screening tool for evaluation of global cognition may be not sensitive to some specific cognition domains, such as spatial localization of a stimulus for goal-directed action.

There are some limitations to be aware of when interpreting our results. First, the CVD cohort was highly selective, and excluded individuals with severe cognitive impairment, significant aphasia, or hemiplegia^23, 24^. Further, 54% of the participants were scored at or above the cutoff on a screening measure of cognitive status (MoCA ≥ 26). Therefore, results may be more drastic if we focus on cohorts entirely with or without cognitive impairment. Second, the suggestion that our results may be related to the possible underlying neural mechanisms for processing of visual perception (neuropsychological outcomes) versus spatial localization (saccadic eye movement), and the underpinning attention systems would be a promising area of future research. In order to better understand those neural networks, imaging technologies should be used to link the identified behavioral markers in the current study with cortical and/or white matter lesions in the relevant pathways.

## Data Availability

All data are available on Brain-CODE.

## Acknowledgements

We would like to thank participants for the time and consent in this study. The authors and investigators would like to acknowledge ONDRI’s project management team, past and present: Susan Boyd, Lisa Desalaiz, Catarina Downey, Stephanie Hetherington, Heather Hink, Ruth Kruger, Sean Lucas, Donna McBain, Andrea Richter, Michelle Vanderspank, Joanna White, and Ashley Wilcox, and the coordinators at each site. The authors would also like to acknowledge the recruitment sites in Ontario, Canada: Baycrest Health Sciences, Toronto; Centre for Addiction and Mental Health, Toronto; Elizabeth Bruyère Hospital, Ottawa; Hamilton General Hospital, Hamilton; Hotel Dieu Hospital, Kingston; London Health Sciences Centre, London; McMaster Medical Centre, Hamilton; Parkwood Institute, London; Providence Care Hospital, Kingston; St. Michael’s Hospital, Toronto; Sunnybrook Health Sciences Centre, Toronto; The Ottawa Hospital, Ottawa; Thunder Bay Regional Health Sciences Centre, Thunder Bay; and Toronto Western Hospital (University Health Network), Toronto.

## Notes

### Competing Interest Statement

The authors have declared no competing interest.

### Funding Statement

This research was conducted with the support of the Ontario Brain Institute, an independent non-profit corporation, funded partially by the Ontario government. Matching funds were provided by participating hospital and research institute foundations, including the Baycrest Foundation, Bruyere Research Institute, Centre for Addiction and Mental Health Foundation, London Health Sciences Foundation, McMaster University Faculty of Health Sciences, Ottawa Brain and Mind Research Institute, Queens University Faculty of Health Sciences, Providence Care (Kingston), St. Michaels Hospital, Sunnybrook Health Sciences Foundation, the Thunder Bay Regional Health Sciences Centre, the University of Ottawa Faculty of Medicine, and the Windsor/Essex County ALS Association. The Temerty Family Foundation provided the major infrastructure matching funds. YC is supported by Indoc. RHS is supported by a Clinician-Scientist Phase II Award from Heart * Stroke. DPM is supported by the Canada Research Chair Program.

### Author Declarations

The research protocol was approved by Research ethics committees at all participating recruitment sites in Ontario, Canada, including Baycrest Health Sciences, Toronto; Centre for Addiction and Mental Health, Toronto; Elizabeth Bruyere Hospital, Ottawa; Hamilton General Hospital, Hamilton; Hotel Dieu Hospital, Kingston; London Health Sciences Centre, London; McMaster Medical Centre, Hamilton; Parkwood Institute, London; Providence Care Hospital, Kingston; St. Michael's Hospital, Toronto; Sunnybrook Health Sciences Centre, Toronto; The Ottawa Hospital, Ottawa; Thunder Bay Regional Health Sciences Centre, Thunder Bay; and Toronto Western Hospital (University Health Network), Toronto.

